# ABO and Rh blood groups, demographics, and comorbidities in COVID-19 related deaths: a retrospective study in Split-Dalmatia County, Croatia

**DOI:** 10.1101/2021.07.16.21260630

**Authors:** Ivan Brdar, Ivan Jerković, Željana Bašić, Nenad Kunac, Deny Anđelinović, Joško Bezić, Ivana Kružić, Arijana Vuko, Šimun Anđelinović

## Abstract

**Aim:** To examine ABO and Rh blood group distribution in COVID-19 related deaths considering demographics and pathological conditions.

**Materials and Methods:** We conducted a retrospective study at the University Hospital Centre Split, Croatia, that included 245 COVID-positive individuals that died from April 8, 2020, to January 25, 2021. From the hospital database, we extracted data on their blood groups, demographics, and pre-existing comorbidities. To compare findings with the general population, we used information from collected blood group donations (n = 101357) and statistical reports of non-COVID deaths from 2019 (n = 4968).

**Results:** The proportion of males was significantly higher in analyzed subjects than in non-COVID deaths from 2019 (63.7% vs. 48.9%, P < 0.001), while the proportion of older individuals did not differ (P = 0.8). The most common pre-existing diseases were hypertension (59.6%), diabetes (37.1%), heart failure (28.8%), digestive disorder (26.5%), and solid tumor (21.6%). The ABO distribution in the deceased and donors’ group showed statistically significant differences, with the higher prevalence of A/AB group and lower prevalence of 0, but with individual differences significant only for AB and non-AB groups. There was a significantly reduced proportion of females within the deceased with group 0 (P = 0.014) and a higher proportion of AB individuals with coronary heart disease (P = 0.024), while other differences were not significant.

**Conclusion:** The study confirmed a higher risk of death in male individuals. The lower proportion of type 0 in deceased individuals was more pronounced in females, implying that group 0 is not necessarily an independent protective factor. Among analyzed comorbidities, coronary heart disease was identified as a potential risk factor for AB individuals.

## 1. INTRODUCTION

Since the outbreak of the COVID-19 in December 2019 and its pandemic spread in the following months, there have been a plethora of studies that aimed to identify potential protective and risk factors and improve the understanding and disease management ^1–4^. Preliminary studies have recognized, and extensive studies that followed proved the association of sex and COVID-19 outcomes, showing that males are more prone to developing more severe disease forms and complications resulting in death ^5,6^. A similar impact was noted for the advanced age, as well as different comorbidities, such as diabetes, hypertension, coronary heart disease, cardiovascular disease, and malignancy, that also showed to increase chances of death ^4,6–9^.

Besides numerous clinical, environmental, and lifestyle factors that have been considered, ABO phenotypes have also been identified to play a possible role in COVID-19 susceptibility and disease severity ^10–12^. Most studies have associated the blood group 0 individuals with a lower risk of the SARS-CoV-2 infection ^10,12–17^, whereas group A ^10,12,14–17^, and in some studies AB group ^17–19^, have been related to the increased risk for contracting SARS-CoV-2 infection and/or disease severity. Although those general findings mainly coincided, the same was not the case for the risk of death in different ABO groups. Zhao et al. ^10^ have associated group 0 with a lower and group A with a higher risk of death. Ray et al. ^20^ also showed a lower risk of death for type O blood group, Takagi ^21^ demonstrated a reduced number of COVID-19-related deaths in populations with the higher prevalence on blood group 0, while Pourali et al. and Zietz et al. ^22,23^ reported increased risk of death in AB group. In contrast, several studies have failed to confirm any association between ABO blood types and COVID-19 related mortality ^16,17,24–27^. Those discrepancies could stem from population differences, type of control group, and many other factors like sex, age, as well as comorbidities that are not always equally considered and controlled for in these types of studies ^25,27,28^.

Since pre-existing comorbidities and ABO phenotypes have been less extensively studied in the COVID-19 deceased individuals, this study aimed to examine differences of the ABO and Rh blood groups distribution in the COVID-19 related deaths and their association to the chronic diseases and individuals’ demographic characteristics. The secondary aim was to reveal potential specificities in the Croatian population, as it has not been regarded in such type of studies.

## 2. MATERIALS AND METHODS

We conducted a retrospective study between March 1, 2021, and May 15, 2021, at the University Hospital Centre Split, Split, Croatia. The study comprised individuals with diagnosed COVID-19 that died in Split-Dalmatia County, Croatia, from April 8, 2020, to January 25, 2021. Using the information from the hospital database, we included only individuals with known ABO blood groups and available information on chronic diseases.

To examine potential demographic differences between the general population of deceased and COVID-19 related deaths, we extracted the data from the statistical report Natural Change in Population for 2019 (n = 4968) ^29^. For blood group distribution in the general population, we used information from collected blood group donations (n = 101357) from January 1, 2016, to April 26, 2021, provided by the Department of Transfusion Medicine of the Split University Hospital Centre, Croatia.

### 2.1 Statistical analysis

Continuous variables were described as mean, standard deviation (SD), and range, while for categorical variables, we provided counts and proportions (%). To examine differences in categorical variables, we used a Pearson’s Chi-squared with Yates’s correction. When test assumptions were not fully met, we approximated P-values using Monte-Carlo simulation with 2000 replicates. All analyses were performed in R statistical software version 3.6.2 and RStudio version 1.2.1335. The statistical significance level was set at P ≤ 0.05.

### 2.2 Ethical approval

The study was approved by the ethical committee of University Hospital Centre Split on June 15, 2020 (500-03/20-01/09; 2181-147-01/06/M.S.-20-12).

## 3. RESULTS

### Participants’ characteristics

From a total of 393 COVID-19 related deaths in the analyzed period, we included 248 patients for which data on ABO blood groups was available. After excluding individuals with no available clinical data in the database (n = 3), the final sample consisted of 245 individuals, 165 (67.3%) male and 80 (32.7%) female subjects. The mean age of male subjects was 74.2 (±11.3), with a range of 30 – 93, while the mean age of female subjects was 78.8 (±10.3) with an age range of 48 – 96.

Sex ratio of the COVID group (2.1:1) statistically significantly differed from non-COVID mortality data from 2019 ^29^ (χ2 = 30.935, P < 0.001), in which proportion of male and female subjects was almost the same (48.9% males and 51.1% females, 0.96:1). When proportion of subjects younger and older than 60 years was considered, no statistically significant differences were noted in either the males (87% vs 86%; χ2 = 0.062, P = 0.803), or the females (95% vs 94%; χ2 = 0.065, P = 0.800).

### Frequency of chronic diseases

Table 1 shows the frequencies of pre-existing chronic diseases in the COVID-19 group. The most common condition in the analyzed individuals was hypertension noted in more than half of subjects (60%) and diabetes identified in 37%. Heart failure, digestive disorder, and solid tumor were recorded for 20-30% of individuals, while other conditions were represented in less than 20% of subjects. Among analyzed conditions, the sex differences were statistically significant only for hypothyroidism (P = 0.047), where a proportion of the disease in female subjects was remarkably higher (64% vs. 36%).

**Table 1.** Frequency of chronic diseases in COVID-19 related deaths.

|  | n (%) | n (%) | n (%) | Sex differences |  |
| --- | --- | --- | --- | --- | --- |
| Disease | Total<br>(n = 245) | Males<br>(n = 165) | Females<br>(n = 80) | $\chi^2$ | P |
| Cerebrovascular disease | 24 (9.8) | 16 (66.7) | 8 (33.3) | < 0.001 | 1.000 |
| Coronary heart disease | 45 (18.4) | 35 (77.8) | 10 (22.2) | 2.177 | 0.140 |
| Heart failure | 69 (28.8) | 48 (69.6) | 21 (30.4) | 0.097 | 0.755 |
| Hypertension | 146 (59.6) | 95 (65.1) | 51 (34.9) | 0.616 | 0.433 |
| Diabetes | 91 (37.1) | 62 (68.1) | 29 (31.9) | 0.004 | 0.952 |
| Digestive disorder | 65 (26.5) | 43 (66.2) | 22 (33.8) | 0.007 | 0.932 |
| COPD | 33 (13.5) | 23 (69.7) | 10 (30.3) | 0.012 | 0.913 |
| Solid tumor | 53 (21.6) | 34 (64.2) | 19 (35.8) | 0.156 | 0.693 |
| Chronic renal disease | 20 (8.2) | 16 (80.0) | 4 (20.0) | 1.021 | 0.312 |
| Hepatitis | 2 (0.8) | 2 (100.0) | 0 (0) | - | - |
| Atrial fibrillation | 41 (16.7) | 30 (73.2) | 11 (26.8) | 0.475 | 0.491 |
| Gout | 19 (7.8) | 15 (78.9) | 4 (21.1) | 0.753 | 0.385 |
| **Hypothyroidism** | 11 (4.5) | 4 (36.4) | **7 (63.6)** | 5.028 | **0.047\*** |
| Rheumatic diseases<sup>†</sup> | 6 (2.5%) | 4 (66.7) | 2 (33.3) | 0.001 | 1.000 |

\*Statistically significant.
† RA, Arthritis, and Morbus Bechterew

### ABO blood group distribution

ABO blood groups distribution showed statistically significant differences in blood donors’ and COVID-19 deceased group (χ2 = 8.204, P = 0.042). However, when blood groups were separately compared (e. g., AB and non-AB group in donors and deceased individuals) the difference was statistically significant only for the AB group (P = 0.034), which was more represented in the deceased individuals. The proportion of Rh-positive and Rh-negative subjects was not significantly different in the donor sample and deceased subjects (P = 0.396).

After considering ABO-Rh blood types, we found no statistically significant differences between groups (χ2= 11.227, P = 0.135). A separate comparison of blood types revealed significant differences for AB- blood type (P = 0.034; Table 3), which was more represented in the group of COVID-positive deceased individuals. Due to the small proportion of Rh- negative individuals, we included only ABO blood groups in further analyses.

**Table 2.** ABO blood group and Rh distribution in donors and deceased subjects.

|  | Blood Group |  |  |  | Rh |  |
| --- | --- | --- | --- | --- | --- | --- |
|  | A | B | AB | O | + | - |
| Donors (n = 101343), % | 39251 (38.7) | 17599 (17.4) | 7046 (7.0) | 37457 (37.0) | 82599 (81.5) | 18754 (18.5) |
| Deceased (n = 245), % | 104 (42.4) | 39 (15.9) | 26 (10.6) | 76 (31.0) | 194 (79.2) | 51 (20.8) |
| $\chi^2$ | 1.274 | 0.262 | 4.507 | 3.447 | 0.720 | |
| P | 0.259 | 0.609 | **0.034\*** | 0.063 | 0.396 |  |

\*Statistically significant.

**Table 3.** ABO-Rh blood group distribution in donors and deceased subjects

|  | **Blood Group** |  |  |  |  |  |  |  |
| --- | --- | --- | --- | --- | --- | --- | --- | --- |
|  | **A+** | **B+** | **AB+** | **0+** | **A-** | **B-** | **AB-** | **0-** |
| **Donors (n = 101343), %** | 31984<br>(31.6) | 14246<br>(14.1) | 5816<br>(5.7) | 30553<br>(30.1) | 7267<br>(7.2) | 3353<br>(3.3) | 1230<br>(1.2) | 6904<br>(6.8) |
| **Deceased (n = 245), %** | 82<br>(33.5) | 31<br>(12.7) | 19<br>(7.8) | 62<br>(25.3) | 22<br>(9.0) | 8<br>(3.3) | 7<br>(2.9) | 14<br>(5.7) |
| **$\chi^2$** | 0.330 | 0.291 | 1.483 | 2.493 | 0.945 | <0.001 | 5.489 | 0.307 |
| **P** | 0.566 | 0.590 | 0.223 | 0.114 | 0.331 | 1.000 | **0.034\*** | 0.579 |

\*Statistically significant.

### Diseases and Blood group distribution

When we analyzed blood groups according to the sex of subjects within the deceased group, overall differences were not statistically significant (Table 4). Nonetheless, when blood groups were separately considered, there was a statistically significantly smaller proportion of females in group 0 (P = 0.014). Among all diseases recorded in the studied subjects, differences in blood group distributions were only statistically significant for coronary heart disease (P = 0.033). This difference was mainly expressed in the AB group, in which more than a third of subjects suffered from coronary heart disease (P = 0.024). Interestingly, the proportion of blood group 0 was twice as higher in participants without disease but with no statistical significance (P = 0.052).

**Table 4.**
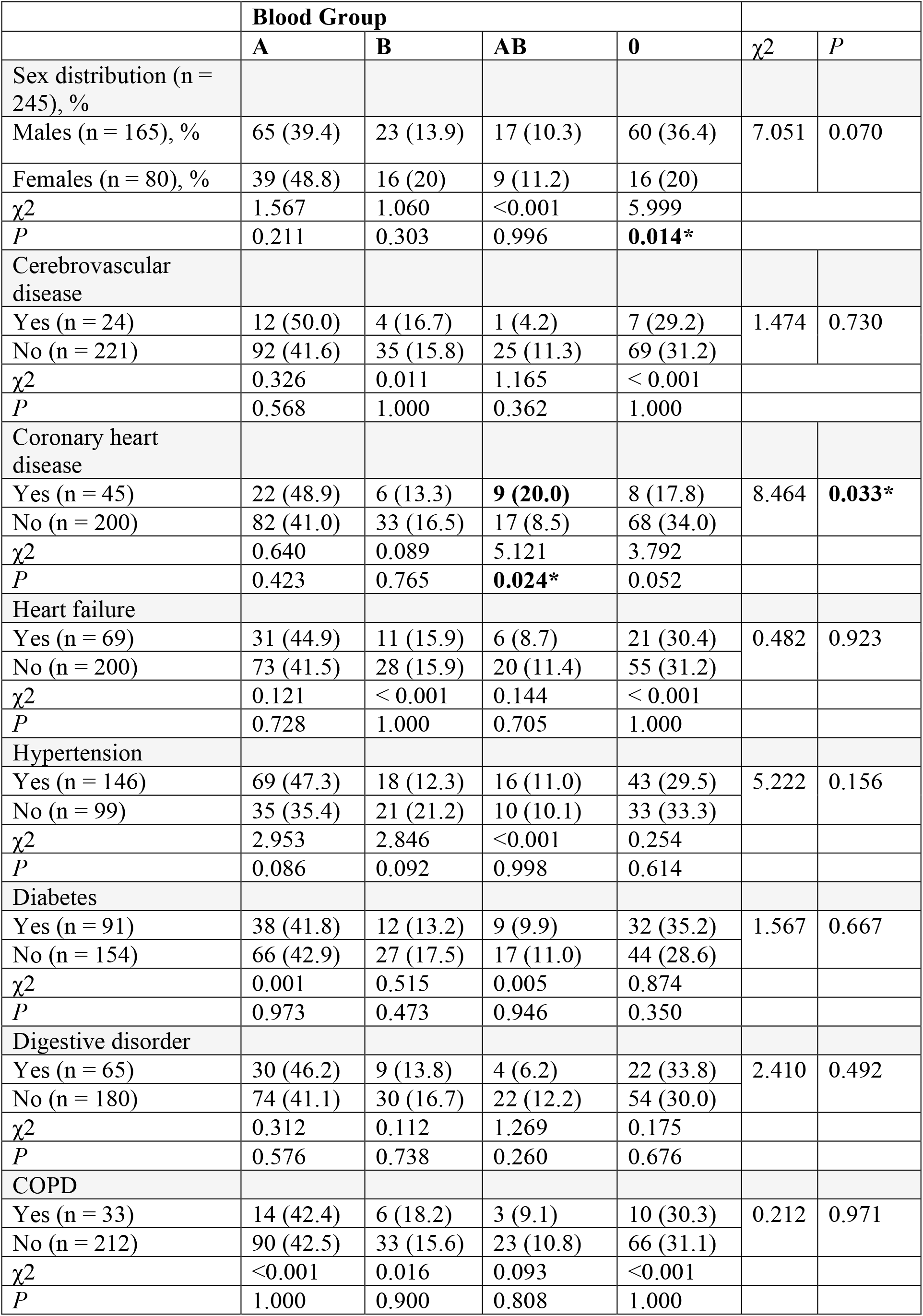
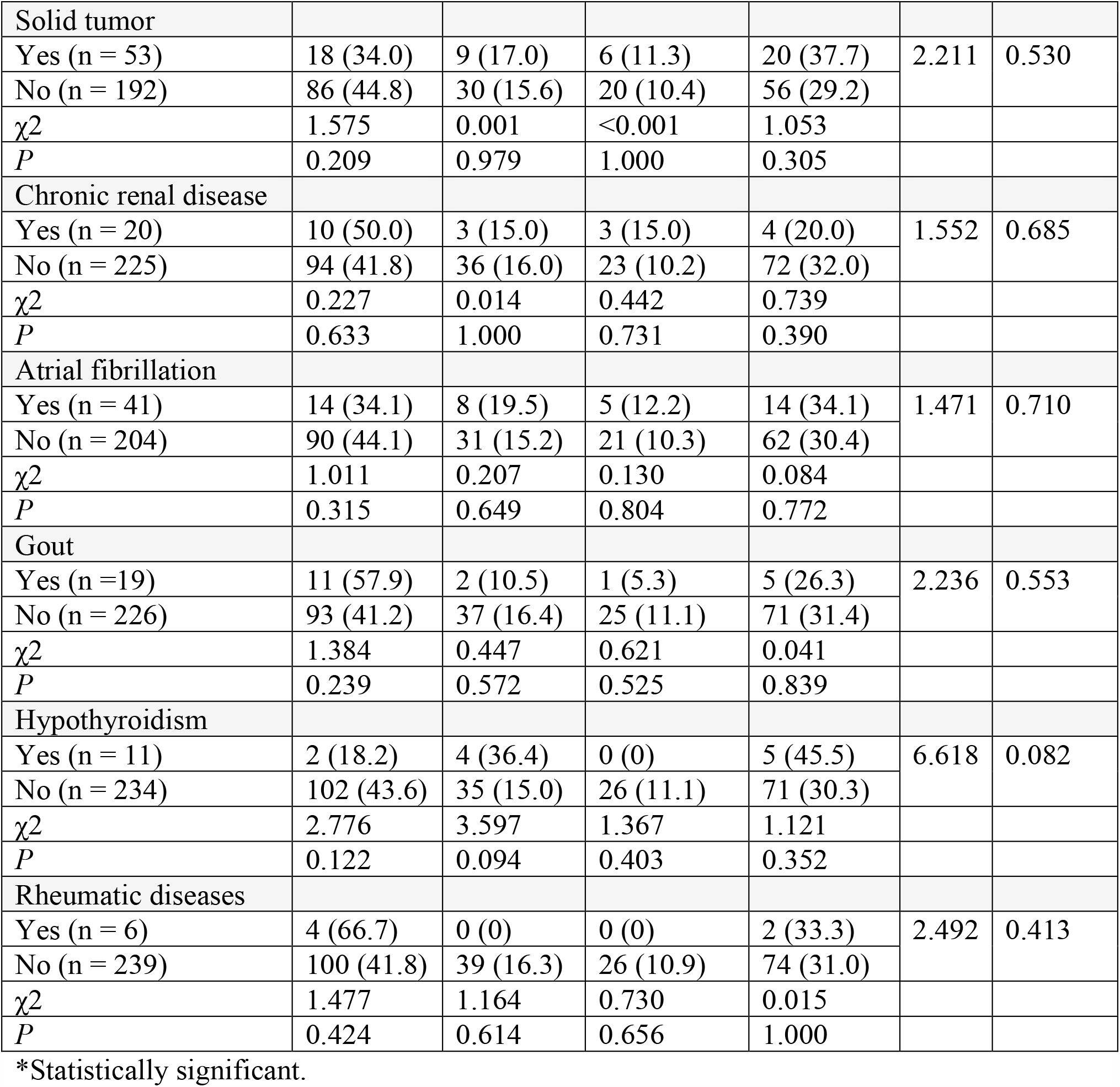
ABO blood group distribution and pre-existing chronic diseases in COVID-19- related deaths

## DISCUSSION

Along with ABO blood groups, this is the first study in the Croatian population that analyzed demographic and pathological conditions in COVID-positive deceased individuals. We showed that the distribution of the ABO blood groups could be associated with the structure of the COVID-related deaths in the analyzed Croatian population sample, but to a limited extent. Blood group AB was significantly more represented in the COVID group than in the group of donors, which is why it might be treated as a potential risk factor in the analyzed population. As expected, blood group 0 was less represented in deceased individuals, but this difference was significant only in females, implying that ABO differences could be masked by the sex of individuals or conditions more frequent in one sex.

Compared to the non-COVID deaths from 2019, where the proportion of males and females was almost equal, the proportion of males was more than twice higher in our study. This result is in line with previous studies that showed significantly higher mortality in male individuals, notwithstanding the approximately equal susceptibility to the disease ^4,5^. Unexpectedly, the proportion of older individuals (>60) was almost equal in COVID and non-COVID-related deaths from 2019, thus not reflecting the increased risk of death for older individuals recognized in the previous studies ^4,7^. The most frequent diseases observed in the analyzed subjects mainly were those previously identified to increase odds of death in COVID-19 cases, such as hypertension, cardiovascular diseases, and diabetes ^7,9,30^. Sex distribution of nearly all diseases did not significantly differ except from hypothyroidism, commonly more frequently diagnosed in females ^31^. Although not statistically significant, coronary heart diseases were almost twice as common in male than female individuals, which was not expected considering that proportion of females that died with diagnosed coronary heart disease in 2019 was slightly higher ^32^. This finding could point out to the additional risk for male individuals with coronary heart diseases.

ABO blood groups distribution showed statistically significant differences in COVID positive deceased individuals and donors’ group, with higher prevalence of A and AB groups and lower prevalence of group 0 in the deceased individuals. Increased proportion of A group and reduced proportion of group 0 in our study was consistent with general findings of disease susceptibility, but also with the disease severity reported in some studies ^10–12,14,16,19^. However, in our study, only AB group was significantly more represented in the group of deceased. This finding is partly in line with meta-analysis that, although statistically non-significant, reported the greatest odds ratio for death in AB group individuals ^23^ and the results reported by Zietz et al. ^22^. One of the possible reasons for this outcome could be biding SARS-CoV-2 to the carbohydrates responsible for the blood group. As those carbohydrates are vastly represented at the respiratory tract mucous membrane, this results in the greatest contact with virus for AB blood type ^23^.

The distribution of Rh+ and Rh blood group in donors and COVID-19 deceased individuals did not differ significantly when analyzed separately neither within ABO Rh groups. This result concurs with the study by Solmaz and Araç ^17^, who found no associations of Rh groups with disease susceptibility, severity or risk of death. However, several studies showed that Rh+ individuals are more likely to test positive ^25^ and associated Rh- individuals with decreased risk for severe disease or death ^20,22^. Due to the mentioned inconsistencies and a limited number of studies, more research is needed to reveal if Rh groups could be related to the risk or resistance to the disease.

We did not find statistically significant differences in sex and ABO blood group distribution within deceased individuals, but when we considered the distribution of 0 and non-0 individuals, there was a significantly lower proportion of females with blood group 0. It is unlikely that this difference is directly linked to the sex and blood group distribution differences, but it might result from sex-specific frequencies of diseases and the blood group-specific susceptibility to a particular disease.

Namely, it has been assumed that anti-A antibodies inhibit the interaction between SARS CoV-1 and ACE2 receptors ^33^; a similar process probably takes place in SARS Co-2 infection^10^. This effect probably also depends on the level of the anti-A antibody ^34^. McVey et al. ^35^showed that among persons with blood group 0, a larger number of women than men have a higher titer of anti-A-antibodies and anti-B antibodies, and a significantly higher number of women have anti-A-antibodies, which corresponds to other similar works. This could be one of the reasons for the lower proportion of deceased females with blood group 0.

Among considered pre-existing chronic conditions, the statistically significant difference in ABO group distributions was observed for coronary heart disease (CHD) that exhibited a higher prevalence of group A and AB and a lower prevalence of group 0. The differences were mostly expressed and statistically significant for AB group, in which more than third of individuals suffered from CHD. This could be explained by generally greater risk in non-O blood type individuals for contracting CHD, particularly group AB, which in some studies showed the highest hazard ratio ^36^. Nonetheless, it is challenging to explain why a similar trend was not observed for heart failure, which exhibited the second smallest differences between ABO blood group distribution according to the χ2 and P-value.

In contrast to the known association between ABO blood groups and CHD, data on the possible association between ABO blood groups and heart failure are lacking. Gotsman et al. ^37^ demonstrated the distribution of blood groups in HF patients similar to that in the general population. The results of this study suggest that the ABO blood group does not play a significant role in the development of HF. However, they show an association between the non-O blood group and poorer prognosis in patients with nonischemic cardiomyopathy. Individuals with non-O blood groups have an increased risk of thrombosis due to elevated levels of von Willebrand factor and factor VIII, with a significantly higher risk of myocardial infarction, peripheral vascular disease, and venous thromboembolism ^38^.

One of the study restrictions is a somewhat smaller sample size, but the major limitation is the unavailability of appropriate control groups that would allow us to draw stronger conclusions. Control groups of COVID-19-negative individuals with data on ABO blood groups and chronic conditions or the same data for individuals that were infected but survived the disease would have enabled more robust analyses regarding the risk factors for disease severity and death in analyzed individuals. For the same reasons, despite sufficient donors’ sample size, we could not exclude the possibility that the general bias of collecting more samples from universal donors (group 0) and less from AB group affected results to some extent. However, since population differences reflect not only in blood types but also in many other genetic and environmental factors, we believe that the present and future studies of this type conducted in various population samples will contribute to creating a clearer and more complete picture of COVID-19 and ABO groups association.

## Data Availability

The data that support the findings of this study are available on request from the corresponding author.

